# Microbial Profile in Bile from Cholecystectomised Patients by Culture and Multiplex PCR

**DOI:** 10.1101/2022.06.29.22277041

**Authors:** Nasreen Farhana, Jannatul Fardows, Mohammad Ashraf Uddin Khan, SM Shamsuzzaman

## Abstract

**Objectives:** Bile in the biliary tract is normally sterile but presence of gallstones, ascending infection from duodenum or bacterial translocation from portal vein leads to microfloral colonization. Therefore, this study was aimed to evaluate the microbiological profile of bile for determination of the appropriate antimicrobials in cholecystectomised patients.

**Design:** This was a descriptive type of cross sectional study from July, 2013 to December, 2014

**Setting:** In a tertiary medical college hospital, Dhaka, Bangladesh

**Participants:** This study included 246 patients irrespective of age and sex who underwent laparoscopic or open cholecystectomy for various indications in a tertiary hospital, Dhaka, Bangladesh.

**Primary and secondary outcome measures:** Similar secondary outcome as in protocol

**Methods:** The intraoperative bile samples were collected and cultured in Blood agar and MacConkey’s agar media aerobically in the laboratory of Microbiology Department, Dhaka Medical College. The isolates were then tested for antibiotic sensitivity pattern. To detect anaerobic bacteria (*Bacteroides fragilis, Clostridium perfringens* and *Fusobacterium nucleatum*), multiplex polymerase chain reaction (PCR) was used with specific primers.

**Results:** Out of 246 bile samples, organisms were identified in 69.51% cases where 48.37% were aerobic bacteria identified by culture and 21.14% were anaerobic bacteria identified by multiplex PCR. *Escherichia coli* (26.61%) were found predominantly followed by *Staphylococcus aureus* (19.35%), *Citrobacter freundii* (14.52%), *Clostridium perfringens* (13.82%), *Pseudomonas* spp. (11.29%), *Salmonella enterica serovar* Typhi 8 (3.45%), and *Acinetobacter* spp. (3.23%). Gram negative bacteria showed resistance to amoxiclav, ceftriaxone and ceftazidime while meropenem, piperacillin-tazobactam and colistin were found more effective antimicrobials.

**Conclusions:** A great proportion of aerobic and anaerobic bacterial infection associated with gallstones or biliary tract obstruction may warrants serious health risk to cholecystectomised patients in this region. Regular surveillance of bile culture from gallbladder should be done after cholecystectomy.

**Strength:**

- The antimicrobial susceptibility pattern of aerobic bacteria was assessed to find out multidrug resistant bacteria
- Multiplex PCR was done to detect important biliary anaerobic bacteria

**Limitations:**

- Isolation by culture and the antimicrobial susceptibility pattern of anaerobic bacteria was not assessed
- Due to lack of primers, PCR could not be done for other biliary anaerobic bacteria, virulence genes and drug resistance genes

**Source(s) of funding for work:** This research received no specific grant from any funding agency in the public, commercial or not-for-profit sectors.

**Competing interests:** None

**Conflict of interests:** None

## Introduction

Bile, a digestive secretion by liver, is concentrated and stored at gallbladder that plays a major role in the digestion of lipids (1). Bile in the gallbladder is usually sterile due to a number of factors including continuous antegrade bile flow toward the duodenum, protective effect of the sphincter of Oddi, bacteriostatic biliary salts and secretory immunoglobulin A (IgA) of the bile (2). The primary mechanism of biliary infection by which microflora enter the bile are thought to be ascending infection from the duodenum due to reflux of contents, blood-borne infection and infection spread through the portal-venous channels and indicate the presence of microflora in 20% - 46% of the patients (3-5). Aerobic bacterial infection is the most common with gram negative preponderance while viral, fungal agents and anaerobic bacteria are uncommon causative agents (4).

Monomicrobial growths are more frequent (30.8% - 96%) in comparison with polymicrobial cultures (4% - 4.8%) (6,7). The most commonly isolated aerobic bacteria from culture of bile have shown principally *Escherichia coli* (17.5% - 62%) (8,9), *Pseudomonas spp*. (5.4% - 23.7%) (6,10), *Acinetobacter* spp. (2.7% - 5.63%) (6,11), *Klebsiella* spp. (5.3% -27%) (4,10), *Staphylococcus aureus* (2.7% - 7.04%) (5,11), *Enteroccocus spp*. (13.1% -15.6%) (4), *Citrobacter* spp. (5.63% - 9.5%) (10,11), *Salmonella enterica* serovar Typhi (2.82% -14%) (7,11). β-glucuronidase producing anaerobes especially *Bacteroides fragilis* (12.86%-58.82%) and *Clostridium perfringens* (32.86% - 36.58%) are recovered in both acutely inflamed and chronically inflamed biliary systems (11,12).

*Salmonella* enterica serovar Typhi is an extreme example of bile-resistant pathogen involving several strategies - invasion of the gallbladder epithelium, escaping from the extremely high concentrations of bile salts and formation of bile inducing biofilms on gallstones in gallbladder lumen (13,14). In general, 2-5% of all individuals who develop clinical or subclinical infection with *S*. Typhi become chronic gallbladder carriers which the development of gallbladder cancer as the worst complication (15,16). The likelihood of carriage is more common in females, increases with age and with gallstones or other gallbladder abnormalities where *Salmonella* form biofilms and continues to intermittently shed organisms for a prolonged period which are generally more numerous in the bile than feces (15,17).

Cholecystectomy does not eliminate the carrier state; hence, the most effective treatment is a combination of surgery and antibiotics. Antimicrobial agents administered empirically should be changed according to their sensitivity suggesting of recent changes in antibiotic-resistant profiles in the patients (16).

To the best of knowledge, no systematic study on bile has been done in Bangladesh to identify aerobic and anaerobic bacteria. So, this study was designed to isolate aerobic bacteria by culture and to see their antibiotic susceptibility pattern along with multiplex PCR was used to detect virulence genes of the anaerobic bacteria in bile.

## Materials and methods

This cross sectional study was conducted in the Department of Microbiology, Dhaka Medical College (located in Dhaka, Bangladesh) from July, 2013 to December, 2014. A total of 246 bile samples of patients irrespective of age and sex underwent open or laparoscopic cholecystectomies were obtained from the Department of Surgery, Dhaka Medical College Hospital, Dhaka. The study was under taken after approval by Research Review Committee (RRC) of the Department of Microbiology of Dhaka Medical College and ethical clearance was taken from the Ethical Review Committee (ERC) of Dhaka Medical College. Informed consents from the patients were obtained before enrolling in the study according to the Helsinki Declaration. Patient’s details such as age, sex, clinical history, symptoms, antibiotic history, ultrasonography reports and information about cholecystectomy operation were extracted from patients’ records using a predesigned data collection sheet.

Non-probability purposive sampling technique were used and the sample size was calculated to 246 by using formula with 20% prevalence of bacteria isolated from the bile at 95% level of confidence with 5% margin of error.

### Patient and Public Involvement

Patients or the public were not involved in the design, or conduct, or reporting, or dissemination plans of our research.

### 2.1 Microbiological examination

#### 2.1.1 Sample collection

About 3 ml of bile was aspirated with a sterile syringe from gallbladder during cholecystectomy and transferred in a sterile falcon tube and carried to the microbiology laboratory of Dhaka Medical College in an hour for immediate inoculation on to 5% sheep blood agar, chocolate agar and MacConkey agar media and incubated at 37°C for aerobic and facultative organisms.

#### 2.1.2 Isolation and identification

The culture media was then examined at 24 and 48 hours for bacterial growth and the isolated bacteria were identified by colony morphology, hemolytic criteria on blood agar media, Gram staining and relevant biochemical tests (18,19).

### 2.2 Antimicrobial susceptibility test

Antimicrobial susceptibility of the isolated bacteria was done by disc diffusion tests using the Kirby-Bauer method using standard antibiotic disks (Oxoid Ltd, UK) (20). Standard antibiotic panel for specific isolated organisms and reading of sensitivity were done according to Clinical and Laboratory Standard Institute (CLSI) guidelines in January 2013 (21).

### 2.3 Extraction of Genomic DNA

Total DNA was extracted from each bile sample by boiling method. About three milliliters of bile were pelleted by centrifugation for 5 min at 14,000g, incubated with 100 µl lytic buffer [composition: 50mM Tris-HCl (pH 8.0), proteinase K (50µg/ml) and 0.5% tween 20 solution] (Promega corporation, USA) for 2 hour at 55°C after vortexing thoroughly and placed in a heat block (DAIHA Scientific, Seoul, Korea) at 100°C for 10 minutes for boiling. After immediately transferred to the ice the mixtures kept for 5 minutes and centrifuged at 10,000 g at 4°C for 15 minutes. The supernatant were used as template DNA and preserved at 4□C for 7-10 days and - 20□C for long time (22).

### 2.4 Amplification of Virulence Genes

Multiplex PCR using specific primer sequences was performed to determine the virulence genes of anaerobic bacteria (*Bacteroides fragilis, Clostridium perfringens* and *Fusobacterium nucleatum*). The primer sequences were for *Bacteroides fragilis* of 230 bp (*Bfr*-forward CTGAACCAGCCAAGTAGCG, *Bfr*-reverse CCGCAAACTTTCACAACTGACTTA), For *Fusobacterium nucleatum* of 161 bp (*nusG* – forward CAACCATTACTTTAACTCTACCATGTTCA, *nusG* – reverse GTTGACTTTACAGAAGGAGATTATGTAAAAATC) and For *Clostridium perfringens* of 1167 bp (CPALPHATOX1-forward AAGATTTGTAAGGCGCTT, CPALPHATOX1-reverse ATTTCCTGAAATCCACTC). PCR was performed in a thermal cycler (Eppendorf AG, Master cycler gradient, Hamburg, Germany) with a final reaction volume of 25µl in a 0.5 mL PCR tube, containing 10 µl of mastermix [premixed mixture of 50 units/ml of Taq DNA polymerase supplied in a proprietary reaction buffer (pH 8.5), 400μM dNTP, 3mM MgCl2 (Promega corporation, USA), 1 µl forward primer and 1 µl reverse primer (Promega corporation, USA), 3 µl extracted DNA and 14 µl nuclease free water for monoplex PCR. After a brief vortex, the PCR tubes were centrifuged in a micro centrifuge machine for few seconds. The PCR consisted of initial denaturation at 95°C for 10 minutes followed by 35 cycles including annealing temperature which varied in different reaction at 52°C (for Bfr), 67°C (for *nus*G) and 46°C (for *cpa-*α) for 45 seconds, elongation at 72°C for 1 min and a final extension at 72°C for 10 minutes was done at the end of the amplification program (23-25).

### 2.5 Analysis of PCR products

The PCR products were analyzed in 1.5% agarose gel electrophoresis followed by staining with ethidium bromide and then observed under UV Transilluminator (Gel Doc, Major science, Taiwan) for DNA bands which were identified according to their molecular size by comparing with 100 bp DNA ladder.

### 2.6 Statistical analysis

Results were presented in the form of tables and figures. The statistical significance was assigned a p value of <0.05 using the z-test of proportion.

## Results

In this study 246 cholecystectomised patients were between the ages of 20 to 70 years and highest 29.68% of them were between 40-50 years of age. Male were found in 64 (26.02%) cases and female were 182 (73.98%) cases and male female ratio was 1:2.84 (Table 1).

**Table 1:** Age and sex distribution of study population (n= 246)

| Age groups<br>(years) | Patients |  | Total<br>n (%) |
| --- | --- | --- | --- |
|  | Male<br>n (%) | Female<br>n (%) |  |
| **20-30** | 8 (18.18) | 36 (81.82) | 44 (17.89) |
| **31-40** | 8 (14.04) | 49 (85.97) | 57 (23.17) |
| **41-50** | 24 (32.88) | 49 (67.12) | 73 (29.68) |
| **51-60** | 12 (29.27) | 29 (70.73) | 41 (16.66) |
| **61-70** | 12 (38.71) | 19 (61.29) | 31 (12.60) |
| **Total** | 64 (26.02) | 182 (73.98) | 246 (100.00) |

\*Female male ratio= 2.84:1

Out of 246 aspirated bile samples, microflora were identified in 171 (69.51%) cases by both culture and multiplex PCR while there was no growth on cultures in 132 (53.66%) cases (Table 2). Single organisms were predominantly isolated in 105 (84.68%) cases while polymicrobial were detected in 19 (15.32%) cases. There was no definite predominance of any combination in polymicrobial infection (Table 3).

**Table 2:** Sex distribution in relation to organisms detected (n=246)

| Sex | No growth<br>on culture<br>n (%) | Detection of positive cases<br>for organism |  | Total positive<br>cases<br>n (%) | p value |
| --- | --- | --- | --- | --- | --- |
|  |  | Aerobic bacteria<br>by culture<br>n (%) | Anaerobic<br>bacteria<br>by PCR*<br>n (%) |  |  |
| **Male**<br>**(n=64)** | 34 (53.13) | 31 (48.44) | 15 (23.44) | 46 (71.88) | p > 0.05 |
| **Female**<br>**(n=182)** | 98 (53.85) | 88 (48.35) | 37 (20.33) | 125 (68.68) |  |
| **Total**<br>**(n=246)** | 132 (53.66) | 119 (48.37) | 52 (21.14) | 171 (69.51) |  |

‘\*’ Anaerobic culture was not done.

**Table 3:** Aerobic organisms isolated from bile of study population (n=119)

| Organism | Culture positive |  | Total<br>n (%) |
| --- | --- | --- | --- |
|  | Monomicrobial<br>n (%) | Polymicrobial<br>n (%) |  |
| *Escherichia coli* | 30 (90.91) | 3 (9.09) | 33 (26.61) |
| *Staphylococcus aureus* | 21 (87.50) | 3 (12.50) | 24 (19.35) |
| *Citrobacter freundii* | 14 (77.77) | 4 (22.22) | 18 (14.52) |
| *Pseudomonas spp.* | 10 (71.43) | 4 (28.57) | 14 (11.29) |
| *Viridans streptococci* | 9 (90.00) | 1 (10.00) | 10 (8.06) |
| *Citrobacter koseri* | 5 (71.43) | 2 (28.57) | 7 (5.65) |
| CONS | 5 (100.00) | 0 (0.00) | 5 (4.03) |
| *Acinetobacter spp.* | 4 (100.00) | 0 (0.00) | 4 (3.23) |
| *Salmonella enterica*<br>serovar *Typhi* | 1 (33.33) | 2 (66.67) | 3 (2.42) |
| *Streptococcus pneumoniae* | 2 (100.00) | 0 (0.00) | 2 (1.61) |
| *Klebsiella pneumoniae* | 2 (100.00) | 0 (0.00) | 2 (1.61) |
| *Klebsiella oxytoca* | 2 (100.00) | 0 (0.00) | 2 (1.61) |
| **Total** | **105 (84.68)** | 19 (15.32) | 124 (100.00) |

\* CONS = Coagulase negative *Staphylococcus*

Of them, aerobic bacteria were isolated by culture in 119 (48.37%) cases and 52 (21.14%) were anaerobic bacteria identified by multiplex PCR and 7(13.46%) anaerobic bacteria were mixed with aerobic bacteria by both culture and multiplex PCR (Table 4).

**Table 4:** Detection of anaerobic bacteria among male and female study population (n=52)

| Sex | Single anaerobic bacteria<br>by PCR<br>n (%) | Anaerobic bacteria and<br>aerobic organisms*<br>n (%) |
| --- | --- | --- |
| Male (n=15) | 13 (86.67) | 2 (13.33) |
| Female (n=37) | 32 (86.49) | 5 (13.51) |
| Total (n=52) | 45 (86.54) | 7 (13.46) |

Note: ‘\*’= anaerobic bacteria detected by PCR and aerobic organisms isolated by culture.

Among isolated organisms, 83 (66.94%) were gram negative bacilli and 41 (33.06%) were gram positive cocci (Figure 1). *Escherichia coli* was predominantly found in 33 (26.61) bile samples whereas *Staphylococcus aureus* were isolated from 24 (19.35%) samples followed by *Citrobacter freundii* in 18 (14.52%), *Pseudomonas* spp. in 14 (11.29%) and *Viridans Streptococcus* in 10 (8.06%). Furthermore low frequencies of *Citrobacter koseri* (7; 5.65%), Coagulase negative *staphylococcus (5; 4*.*03%), Acinetobacter* spp. *(4; 3*.*23%), Salmonella enterica serovar Typhi (3; 2*.*42%), Streptococcus pneumoniae (2; 1*.*61%)* and *Klebsiella* spp. (2; 1.75%) were also isolated (Table 3). *Clostridium perfringens, Fusobacterium nucleatum* and *Bacteroides fragilis* were detected in 34 (13.82%), 10 (4.07%) and 8 (3.25%) cases respectively by multiplex PCR (Table 5) (Figure 2,3).

**Figure 1:**
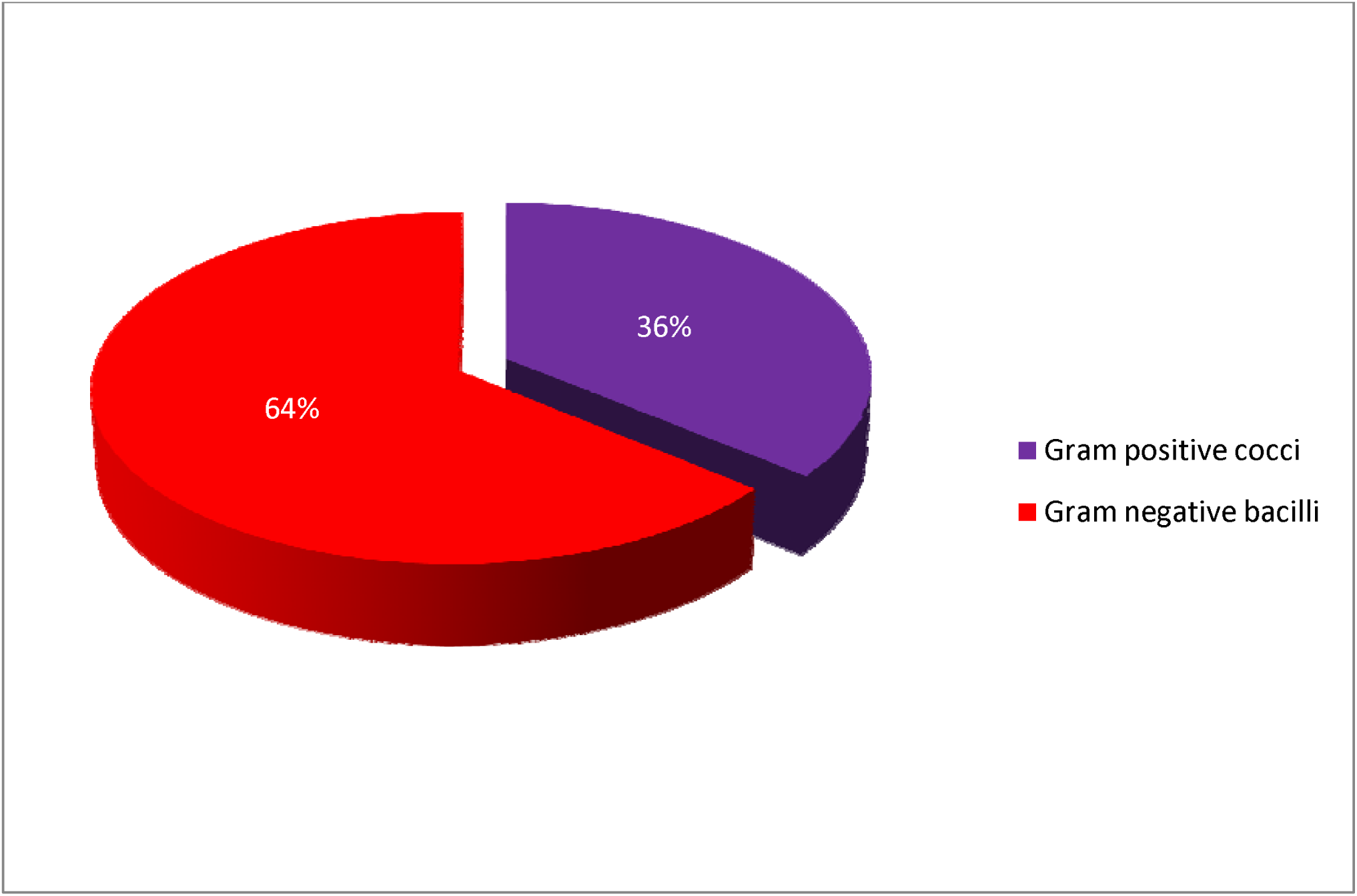
Organisms grown in bile culture

**Table 5:** Distribution of anaerobic bacteria identified by multiplex PCR from bile (n=246)

| Organisms | Number identified by PCR<br>n (%) |
| --- | --- |
| *Clostridium perfringens* | 34 (13.82) |
| *Fusobacterium nucleatum* | 10 (4.07) |
| *Bacteroides fragilis* | 8 (3.25) |

**Figure 2:**
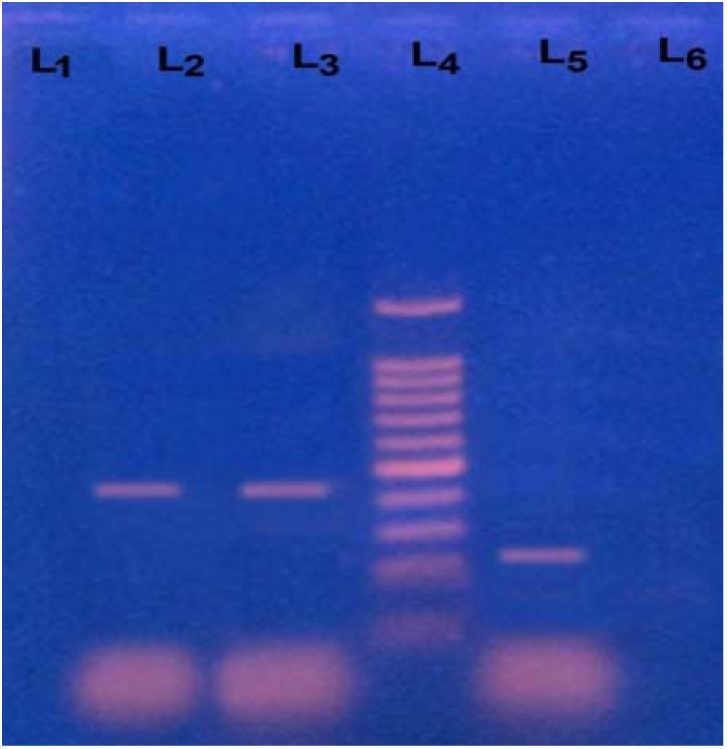
The photograph showing of amplified DNA of *Bacteroides fragilis*, here in lane 2: negative control (DNA of *Pseudomonas aeruginosa*), in lane 4: 100bp DNA ladder and in Lane 5: bile sample (positive for *Bacteroides fragilis* of 230 bp)

**Figure 3:**
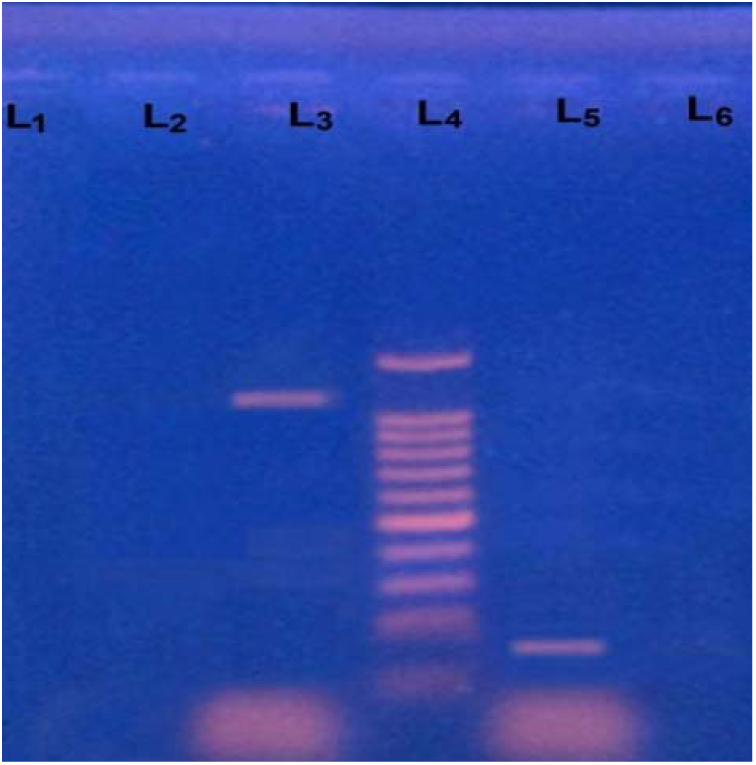
The photograph showing amplified DNA of *Clostridium perfringens* and *Fusobacterium nucleatum*, here in lane 1: negative control (DNA of Fusobacterium necrophorum), in lane 2: bile sample (positive for *Clostridium perfringens* of 1167 bp), in lane 4: 100bp DNA ladder and in lane 5: bile sample (positive for *Fusobacterium nucleatum* of 161 bp)

According to clinical profiles of 246 the study population, the third generation cephalosporins were used as preoperative prophylactics (Table 6) but antibiotic sensitivity tests of isolated bacteria showed that third generation cephalosporins were most resistant antibiotics for gram negative bacteria while carbapenems and colistin showed most susceptible antibiotics. On the other hand, *Salmonella* Typhi and *Staphylococcus aureus* were found most sensitive to ceftriaxone and ciprofloxacin (Table7).

**Table 6:** Clinical profile and other characteristics of the study population (n=246)

| **Clinical profile of study population** | **Frequency**<br>**n (%)** |
| --- | --- |
| **A) Clinical presentation** |  |
| Pain in the abdomen | 246 (100.00) |
| Nausea, vomiting, dyspepsia | 74 (30.08) |
| Intolerance to fatty food | 50 (20.33) |
| Fever | 82 (33.33) |
| Jaundice | 7 (2.85) |
| **B) Co-morbidity** |  |
| Hypertension | 23 (9.35) |
| Diabetes mellitus | 21 (8.54) |
| Others (specifically related to cholecystitis) | 12 (4.88) |
| **C) Clinical diagnosis** |  |
| Acute on chronic cholecystitis | 15 (6.09) |
| Chronic cholecystitis | 34 (13.82) |
| Cholelithiasis / Calculus cholecystitis | 190 (77.24) |
| Choledocholithiasis | 7 (2.85) |
| **D) Types of surgery** |  |
| Open cholecystectomy | 21 (8.45) |
| Laparoscopic cholecystectomy | 225 (91.55) |
| **E) Preoperative antimicrobials used** |  |
| Ceftriaxone | 131 (53.25) |
| Cefuroxime | 46 (18.70) |
| Ceftriaxone and metronidazole | 36 (14.63) |
| Cefixime | 23 (9.35) |
| Cefixime and metronidazole | 7 (2.85) |
| Ciprofloxacin | 3 (1.22) |

**Table 7:**
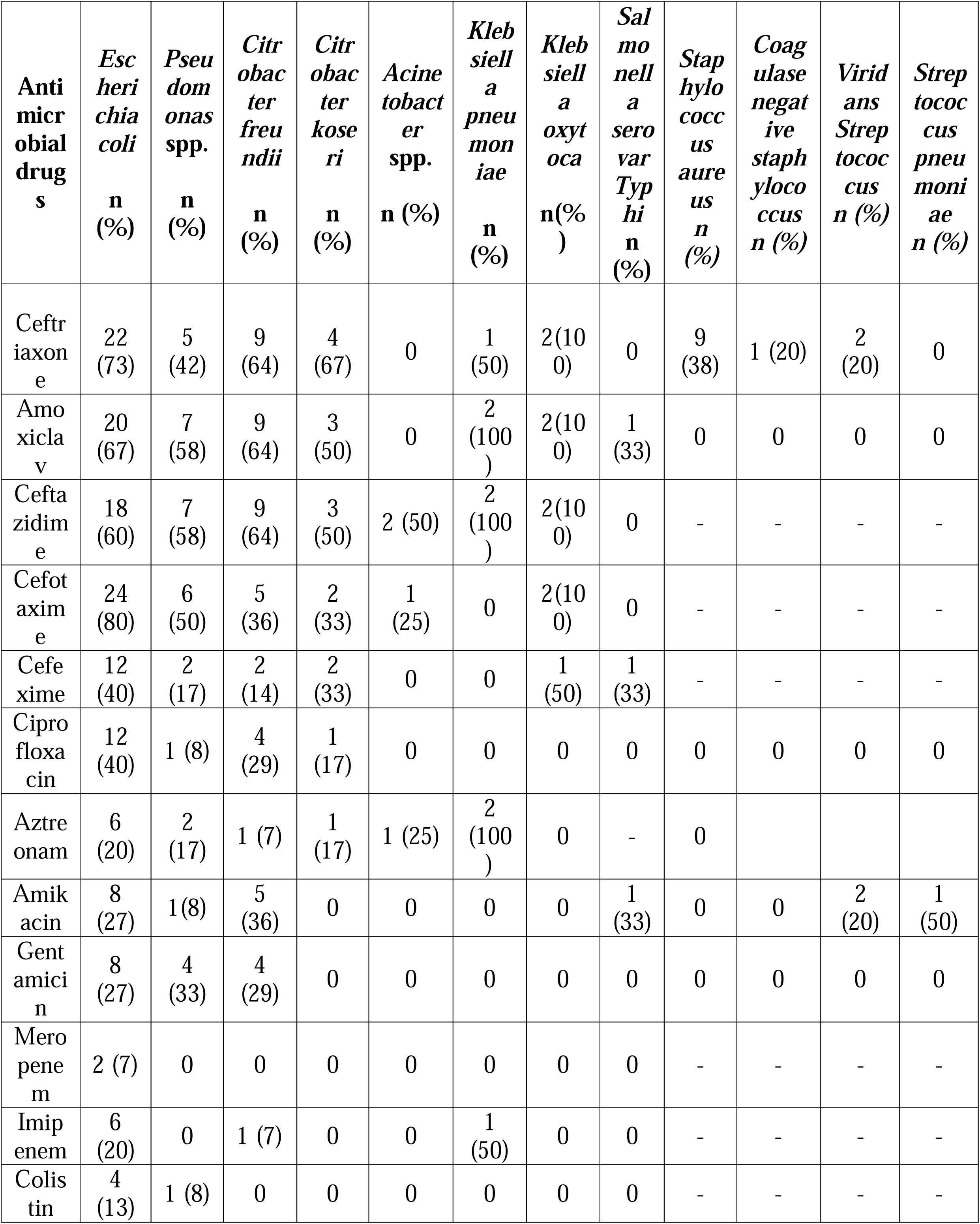
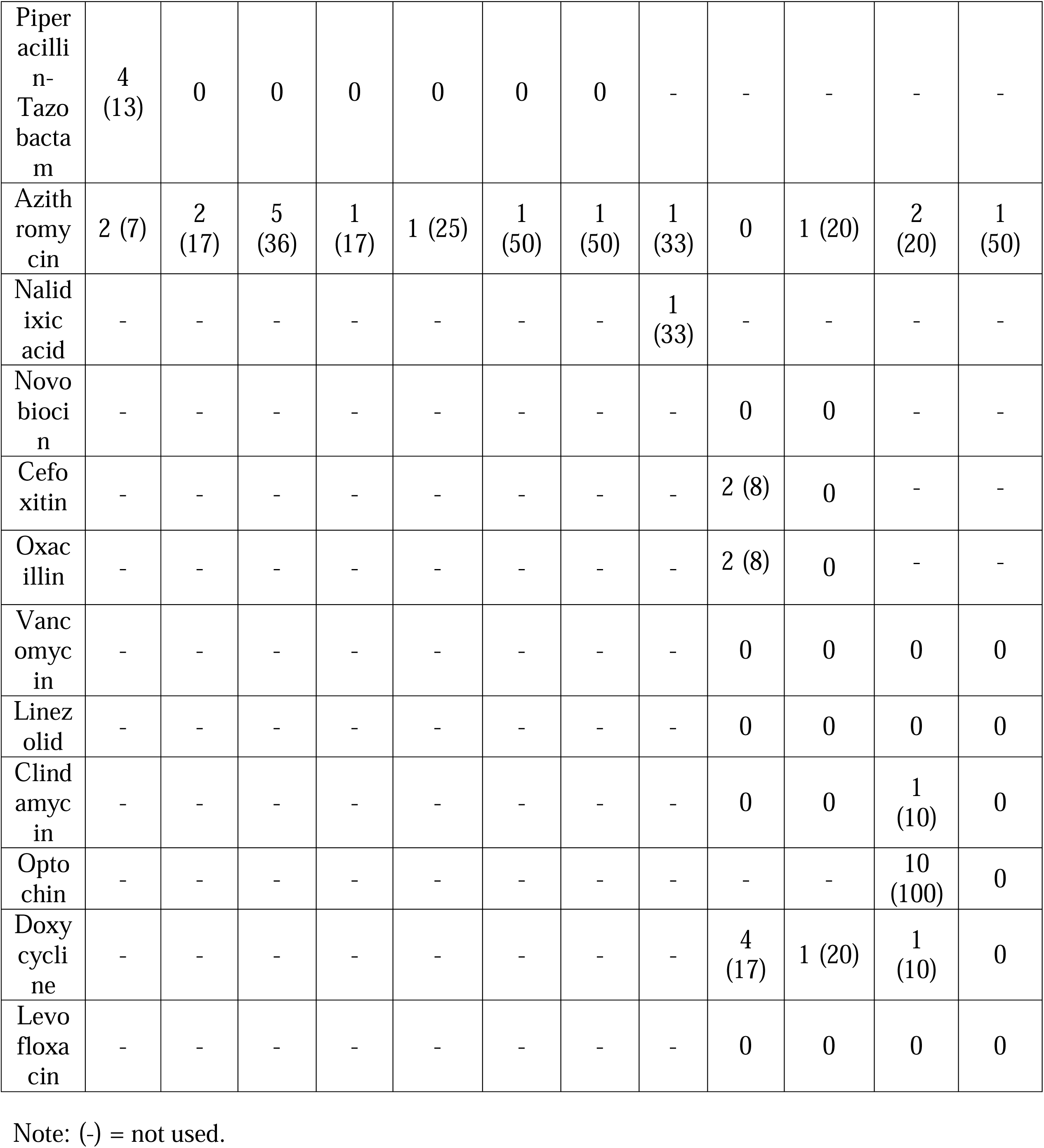
Antibiotic resistance pattern of isolated organisms to different antibiotics (n=105)

## Discussion

Biliary microflora is important agents in the gallbladder when biliary obstructions are often due to gallstones (5). Therefore this study was to identify the aerobic microbial flora by culture along with antibiotic sensitivity and detection of anaerobic bacteria in the bile by multiplex PCR.

While reviewing the literature, microflora were identified in other studies with the rates of 26-58% which was comparable to this study (4,7,9,27). In a study by Dongol et al. showed a lower proportion (20%) of positive bile cultures while Ahmad et al. has reported considerably higher (58.58%) than the present study (9,16). Mixed aerobes and anaerobes were identified in this study by both culture and multiplex PCR in 7 (13.46%) cases while some studies found in 5-6% cases from bile cultures (6,8). The increased rate of mixed growth may be due to some antibiotics resistance bacteria along with food habits and environmental factors.

β-glucuronidase catalyse the hydrolysis of bilirubin glucuronide to form insoluble salts that plays a role in gallstone formation. Among anaerobic bacteria, β-glucuronidase producing *Clostridium perfringens* and *Bacteroides fragilis* were the most frequent isolates which was also supported by Atia et al. (25). *Escherichia coli* has been also reported as the predominant flora by other authors and attributed to the activity of its β glucuronidase enzyme in formation of calcium billirubinate gallstone formation (4,10,16,26).

In this study, *Salmonella* Typhi was isolated in 3.25% while similar prevalence of invasive *S*. Typhi in bile culture was found in a study by Dongol et al. (16). *S*.Typhi was also found at a higher proportions from bile samples (8-14%) due to endemic typhoid fever in other studies (4,13,28). There was also a low prevalence (0.41%, 4/977) for *Salmonella* in bile cultures in case-control study by Safaeian et al.(29).

Among the isolated gram-positive cocci, *Staphylococcus aureus* was found in 24 (19.35%) cases in this study which correlates with the study by Suri et al.(30). Low isolation of *S. aureus* was reported in 0.8%-5.6% bile in some studies while Fatemi et al. reported higher (29.67%) *S. aureus* than the present study which might be due to geographical variation and antibiotic policy of the respective country (7,23,25).

Sensitivity to third- and fourth-generation cephalosporins in antimicrobial susceptibility test of isolated gram negative bacilli was higher as compared to aminoglycoside in some studies while Shahi et al. observed high level of resistance to third generation cephalosporins which is similar to the present study (7,12,28). Meropenem, piperacillin-tazobactam and colistin showed good sensitivity against isolated organisms and may be used as the first line of preoperative prophylaxis in cholecystectomy. The most high resistance profiles in this study were *Escherichia coli, Pseudomonas aeruginosa, Klebsiella spp*. and *Citrobacter freundii* similar to the study by Kaya et al. (7). S. Typhi was found most susceptible to ciprofloxacin and ceftriaxone while another study by Dongol et al. had showed resistance to only nalidixic acid in *Salmonella* from bile (16). In the present study, *Staphylococcus aureus*, Coagulase negative *Staphylococcus, Viridans streptococci* and *Streptococcus pneumoniae* were found highly resistant to ceftriaxone, doxycycline, oxacillin and cefoxitin which was supported by Ahmad et al.(9).

In a number of studies revealed that incidence of biliary obstruction due to gallstones is common above the age of forty in 25-53% patients which also included the findings of this study (4,7,10, 23). Predominance of female patients were found in this study and attributed to estrogens with HMG-Co-A reductase enzyme causing increased formation of cholesterol stones (25).

## Data Availability

All data produced in the present work are contained in the manuscript

## Acknowledgments

We acknowledge the supports of the Surgery Departments of Dhaka Medical College Hospital, Dhaka, Bangladesh for providing the opportunity and resources to undertake the study.

## Author statements

**Nasreen Farhana:** Conceptualization, Methodology, Formal analysis, Investigation, Writing-Original draft preparation

**Jannatul Fardows:** Data curation, Visualization, Writing - Review & Editing

**Mohammad Ashraf Uddin Khan:** Resources, Visualization

**SM Shamsuzzaman:** Supervision, Project administration, Writing - Review & Editing

**Data statement:** technical appendix

